# Sustained attention processes in very preterm adolescents and their relationship to socio-emotional competence

**DOI:** 10.1101/2024.09.30.24314596

**Authors:** NB. Fernandez, V. Siffredi, J. Awada, J. Miehlbradt, C. Borradori-Tolsa, MC. Liverani, R. Ha-Vinh Leuchter

**Affiliations:** Division of Development and Growth, Department of Paediatrics, Gynaecology and Obstetrics, Geneva University Hospitals, Geneva, Switzerland; Institute of Bioengineering, Center for Neuroprosthetics, Ecole Polytechnique Fédérale de Lausanne, Switzerland; Department of Radiology and Medical Informatics, Faculty of Medicine, University of Geneva, Switzerland; SensoriMotor, Affective and Social Development Laboratory, Faculty of Psychology and Educational Sciences, University of Geneva, Geneva, Switzerland

**Keywords:** **Keyword:** preterm, prematurity, sustained attention, socio-emotional functions, fMRI, gradCPT

## Abstract

Very preterm (VPT) adolescents are at high risk of impaired sustained attention processes, as well as behavioral and socio-emotional problems. Previous studies have highlighted altered attentional patterns of brain activation in this population, but results are inconsistent. The current study aims to explore brain activity related to sustained attention in VPT and full-term adolescents aged 11-18, as well as its associations with attentional capacities and socio-emotional competences. Event-related functional MRI (fMRI) was used to assess sustained attention performance and associated brain activations by comparing VPT (n = 34) and their age-matched full-term (FT, n = 28) peers from a previously validated continuous performance task with gradual onset (gradCPT) paradigm, using two different modality versions (i.e., face and scene). In both groups, linear regression analyses were performed to examine associations between attentional and socio-emotional difficulties and brain activations related to sustained attention. Results show preserved sustained attention processes in VPT adolescents, indicated by comparable behavioral attentional performance and cerebral patterns of activations in both groups across the two modalities of the gradCPT. In addition, VPT adolescents showed over-recruitments in posterior occipital areas compared to FT adolescents. Moreover, higher socio-emotional difficulties (i.e., higher anxiety and social difficulties) in VPT were linked to altered activations specifically in the right middle frontal gyrus, occipito-temporal gyri and bilateral cerebellum, but exclusively observed during the face modality of the gradCPT. Overall, these results suggest that despite preserved sustained attention competences, VPT adolescents present a less mature sustained attention cerebral network, particularly during a task with a social context.

## 1. Introduction

Attentional processes are crucial for development and underpin multiple cognitive functions essential for daily life activities (Ruff and Rothbart 2001, Reynolds and Romano 2016, Fortenbaugh, DeGutis et al. 2017, Fisher 2019). Among these processes, sustained attention stands out as the ability to maintain sensitivity for upcoming stimuli over an extended period of time. Categorized as a fundamental sub-component of attention in theoretical frameworks (Sohlberg and Mateer 2001), sustained attention is closely interconnected to selective attention and inhibition in daily-life, as sensory stimuli need to be selectively processed while excluding others for a continuous amount of time. In laboratory settings, sustained attention (also known as vigilance or alertness over time) has been most frequently studied using non-X continuous performance tasks (CPTs), for instance Conner’s CPT (Conners, Epstein et al. 2003), sustained attention to response task (SART; Robertson, Manly et al. 1997), or the gradual-onset CPT (gradCPT; Esterman, Noonan et al. 2013, Rosenberg, Noonan et al. 2013, Rosenberg, Finn et al. 2015, Rosenberg, Finn et al. 2016), which involves continuous discrimination of visual stimuli presented in a constant stream. The gradCPT, in particular, is a novel short-duration non-X CPT paradigm characterized by a gradual transition between two types of stimuli and requiring frequent overt responses to nontarget stimuli, while responses to rare target stimuli need to be inhibited. Contrasting the two types of conditions (nontarget and target) allows to assess behavioral attentional performance as well as the cerebral areas associated with the sustained attention processes. These processes are supported by the overlap of distinct attentional networks, encompassing the fronto-parietal network and the cingulo-opercular network (Menon and Uddin 2010, Petersen and Posner 2012, Sadaghiani and D’Esposito 2015). In particular, previous neuroimaging works in healthy adults reported the engagement of bilateral, predominantly right-dominant, fronto-parieto-occipital areas, especially in frontal cortices (i.e., pre-supplementary motor area, medial prefrontal cortex, midcingulate cortex, inferior frontal junction, ventral premotor cortex), parietal cortices (i.e., superior and inferior parietal lobes, intraparietal sulcus, temporo-parietal junction), the temporo-occipital junction, the anterior insula, the thalamus, and the cerebellum (Langner and Eickhoff 2013, Fortenbaugh, DeGutis et al. 2017, Fisher 2019). In parallel, a comparable sustained attention network has been highlighted in typically developing children and adolescents from 8 to 18 years (Morandini, Silk et al. 2020), although this younger population tend to exhibit lower top-down (i.e., frontal and subcortical areas) and enhanced bottom-up sensory stimulus processing (i.e., posterior occipito-temporal areas) compared to adults (Rubia 2013, Fortenbaugh, DeGutis et al. 2017). These findings coincide with evidence demonstrating that sustained attention continues to mature through adolescence in typical development (Constantinidis and Luna 2019). While neural correlates of attention networks are now better understood in neurotypical children and adolescents (Posner, Rothbart et al. 2016), less is known about the neural substrates underlying sustained attentional functions in populations at risk for attentional difficulties.

Notably, the preterm-born population (< 32 completed weeks of gestation) faces an increased risk of impairments in attentional processes, including selective and sustained attention (Aarnoudse-Moens, Weisglas-Kuperus et al. 2009, Mulder, Pitchford et al. 2009), which are crucial for everyday goal-directed adaptative behavior, as well as in socio-emotional competences (Montagna and Nosarti 2016) and affective behavior (Hornman, De Winter et al. 2016). In the very preterm (VPT) population, these difficulties, which have been documented throughout childhood and adolescence, impact not only academic achievement (Urben, Van Hanswijck De Jonge et al. 2017) but also the establishment of peer relationships and daily functioning (Montagna and Nosarti 2016). Importantly, attentional difficulties tend to persist into adulthood, consequently reducing the quality of life in this population (Linsell, Johnson et al. 2019). In preterm adolescents and adults, previous evidence has linked attentional impairments to structural cerebral alterations, such as lower cortical densities (Urben, Van Hanswijck De Jonge et al. 2017), as well as functional cerebral alteration with changes in functional connectivity (Wheelock, Lean et al. 2021) and modulations in cerebral recruitment in regions supporting attentional performance (Hadaya and Nosarti 2020). More particularly, task-based studies have associated selective and sustained attention processes with a complex pattern of alterations in the fronto-striatal networks in preterm adolescents and young adults compared to full-term (FT) controls (Nosarti, Rubia et al. 2006, Lawrence, Rubia et al. 2009, Nosarti, Shergill et al. 2009). Nevertheless, these studies show heterogeneous results. On one hand, Nosarti, Rubia et al. (2006) reported a decrease in activation in prefrontal regions (i.e., IFG, anterior cingulate cortex), subcortical areas (i.e., caudate nucleus, thalamus) and the cerebellum, as well as hyper-activations in prefrontal regions (i.e., dorsolateral and orbitofrontal prefrontal cortices), in the insula, and in temporo-occipital regions in young adolescents. On the other hand, Lawrence, Rubia et al. (2009) only reported an increase in activation in posterior cerebral areas (i.e., middle temporal gyrus, posterior cingulate cortex and the precuneus) in VPT adults compared to their FT counterparts. These changes in activation were interpreted as a reorganization of the attentional network into a more effective pathway to compensate for potential cerebral alterations (Réveillon, Hüppi et al. 2018). However, authors also suggested an intact organization of the attentional network in VPT individuals as they reported a broad overlap of fronto-cingulo-parietal activations between VPT and FT young adults, with no significant between-group differences (Daamen, Bäuml et al. 2015). Despite the growing interest in characterizing attentional deficits related to prematurity, complementary studies are required to unravel brain correlates underlying sustained attention in this population.

In parallel to attentional problems, deficits in socio-emotional processes in VPT children and adolescents have been linked to atypical structural (Fischi-Gómez, Vasung et al. 2015, Siffredi, Liverani et al. 2023) and functional (Papini, White et al. 2016, Siffredi, Liverani et al. 2023) connectivity, as well as to alterations in functional brain patterns observed using a task-based fMRI paradigm (Urbain, Sato et al. 2019). This latter study, in which participants were presented with faces depicting various emotions that they had to inhibit, demonstrated a decrease in brain activity in a right-lateralized fronto-parietal network during emotion regulation. Moreover, prior behavioral work highlighted impaired cognitive control (i.e., the ability to maintain information and inhibit irrelevant information) in VPT adolescents with social weaknesses (Twilhaar, de Kieviet et al. 2019). However, to our knowledge, there is currently no neuroimaging study linking atypical socio-emotional abilities to specific prematurity-related alterations in the attentional network in VPT adolescents. Given that social difficulties might arise from impaired interactions between cognitive control systems and emotion processing, and recognizing the importance of socio-emotional competences in daily life activities, it is crucial to explore the neuronal pathways related to sustained attention and socio-emotional deficits in a population of VPT adolescents.

Hence, the overall objective of the present study is to enhance understanding of cognitive and emotional impairments associated with prematurity, more particularly by linking attentional and socio-emotional abilities to attentional brain networks in VPT adolescents. To this end, we used a previously validated fMRI gradCPT paradigm, which offers several advantages (Esterman, Noonan et al. 2013, Rosenberg, Finn et al. 2016). First, this paradigm enables fine-grained investigations of intra-individual reaction time variability, a robust indicator of attentional performance (Hylan 1898, Rosenberg, Noonan et al. 2013, Rosenberg, Finn et al. 2015), which has previously been related to impairments in executive functions and attention (Bellgrove, Hester et al. 2004). Second, the introduction of gradual transitions between stimuli removes abrupt visual onsets of trials, known to exogenously capture attention (Yantis and Jonides 1984), and thereby increases demands for maintaining sustained attention throughout the task (Rosenberg, Noonan et al. 2013). This results in a shorter task duration compared to others CPTs. Third, this task has been more recently used to characterize sustained attention in neurotypical and clinical populations, including children and adolescents presenting attention deficit hyperactivity disorder (ADHD) (Rosenberg, Finn et al. 2016), but has never been applied in the VPT population. Therefore, the unique features of this novel CPT paradigm offers valuable insight into evaluating cerebral engagement underlying sustained attention, assessed via response inhibition process, in VPT adolescents.

In the current work, we initially investigated potential sustained attention difficulties related to prematurity by comparing VPT and FT adolescents based on their behavioral performance and corresponding cerebral activation patterns assessed using whole-brain fMRI analyses during conditions requiring response inhibition. Subsequently, we explored the influence of both individuals’ sustained attention performance and socio-emotional abilities on the brain systems governing sustained attention processes. Drawing upon previous evidence, we postulate that VPT adolescents, especially those presenting socio-emotional difficulties related to prematurity, may exhibit differential engagements in sustained attention networks compared with FT adolescents. Specifically, we expected to observe a decrease in activation in fronto-parietal areas associated with top-down mechanisms, alongside an increase in activation in bottom-up posterior visual areas.

## 2. Materials and Methods

### 2.1 Participants

One hundred and sixty-five VPT adolescents were invited to participate in the “Mindful preterm teens – follow up” study between January 2021 and August 2022. 42 adolescents accepted to participate and were enrolled in the study. All VPT were born before 32 gestational weeks between 01.01.2003 and 31.12.2008 in the Neonatal Unit at the Geneva University Hospital, Switzerland, and were followed by the Division of Child Development and Growth at the Geneva University Hospital. Participants were excluded if they had an intelligence quotient below 70, any sensory or physical disabilities (blindness, hearing loss, cerebral palsy), or an insufficient understanding of French. Additionally, 29 term-born typically developing adolescents born after 37 gestational weeks between 01.01.2003 and 31.12.2008 and matching for age were recruited through the community and were enrolled in the current study as FT control participants. The Swiss Ethics Committees on research involving humans approved the all experimental protocols (ID: 215-00175). Written informed consent in accordance with regulations of the ethic committee at the University Hospital of Geneva was obtained from the principal caregiver and from the participant. All participants received a gift voucher of 50 Swiss francs for their participation in the study.

### 2.2 Neonatal/Demographic characteristics and Neurobehavioral measures

Neonatal characteristics were retrieved from medical records. Demographic measures (i.e., socio-economic status) as well as general intellectual abilities and neurobehavioral measures (i.e., executive, attentional, behavioral and socio-emotional functioning) were assessed in both VPT and FT participants using neuropsychological testing, self-reported and parent-reported questionnaires.

#### (i) Socio-economic status

Socio-economic status of the parents was evaluated based on parent-reported demographic questionnaires and using the Largo scale. This validated 12-point score scale is based on maternal education and paternal occupation and ranges from 2 to 12 points. Higher scores indicate lower socio-economic level (Largo, Pfister et al. 1989).

#### (ii) General intellectual abilities

The General Ability Index (GAI) was assessed using the Wechsler Intelligence Scale for Children-5th Edition (WISC-V) to evaluate the participants’ general intellectual functioning (Wechsler 2014). The GAI is derived from the core verbal comprehension and perceptual reasoning subtests. The GAI has a mean of 100 and a standard deviation of 15.

#### (iii) Executive and attentional measures

The parent-rated version of the Behavior Rating Inventory of Executive Function (BRIEF; Gioia, Isquith et al. 2000) was used to assess adolescents’ executive functions in everyday life. Scored on a 3-point Likert scale, it comprises 86 items over 8 executive functioning scales which are categorized into two standardized subscales, i.e., the Behavioral Regulation Index (BRI) encompassing Inhibit, Shift and Emotional control scales, as well as the Metacognition Index (MI) encompassing Initiate, Working Memory, Plan/Organize, Organization of Materials and Monitor scales. Taken together, these two indices form the Global Executive Composite summary score (GEC). In the present study, we analyzed T-scores of the two indices (BRI and MI) and of the composite score (GEC). Lower scores indicate better executive functioning.

The adolescents’ attentional abilities were firstly assessed using the French version of the self-reported Conners’ Rating Scale Third Edition (Conners 3-SR)(Conners 2008; Conners 3rd edition: Manual. Toronto, Ontario, Canada). With 97 items scored on a 4-point Likert, the Conners 3-SR comprises five content scales. Inattention and Hyperactivity/Impulsivity scales represent the two primary ADHD scales, while Learning Problems, Defiance/Aggression and Family Relations scales are indices of common difficulties that are likely to co-exist with ADHD symptoms. Additionally, the ADHD Index can be computed using 10 over the total of 97 items. In the present study, we focused on T-scores for Inattention, and Hyperactivity/Impulsivity scales as well as the ADHD index. Higher scores indicate increased attentional difficulties.

Attentional abilities were secondly assessed using computerized tasks from the Test of Attentional Performance (TAP; Zimmermann and Fimm 1994, Zimmermann and Fimm 2004)(version 2.3.1., 2017). Inhibitory control was evaluated with two versions of the Go/No-Go paradigm (1 target amongst 3 stimuli or 2 targets amongst 5 stimuli), based on T-scores for the number of errors. These scores are considered valuable indicators of impulsivity and control deficits (Zimmermann and Fimm 1995).

#### Socio-emotional measures

Adolescents’ socio-emotional abilities in daily life were assessed using the parent version of the Strength and Difficulties Questionnaire (SDQ) (Goodman 1997, Goodman 2001). Scored on a 3-point Likert scale, the SDQ parent questionnaire consists of 25 statements and evaluates five subscales, i.e., Emotional problems, Conduct problems, Hyperactivity, Peer problems and Prosocial scales. The present study focusses on scores of the three socio-emotional individual scales: i. Emotional problems, ii. Peer problems, and iii. Prosocial scales. Higher scores indicate increased difficulties for Emotional and Peer problems scales, while higher scores in the Prosocial scale indicate better prosocial skills. The presence and severity of anxiety symptoms in adolescents were assessed using a French version of the Multidimensional Anxiety Scale for Children 2^nd^ Edition – Self-Report (MASC; March 1997). The MASC consists of 39 items scored on a 4-point Likert scale and gives a Total Anxiety score (based on the sum of all 39 items). Higher scores indicate a higher level of anxiety.

### 2.3 Neonatal/Demographic characteristics and Neurobehavioral measures analysis

Statistical analyses of participants’ characteristics and neurobehavioral measures were performed using the R Software (version 4.1.2, R.C. Team, 2021) and R Studio (version 2022.02.0). Comparisons of the VPT and the FT groups regarding their neonatal and demographic characteristics (i.e., gestational age, birth weight, age at assessment, socio-economic status, and GAI) as well as outcome measures of neurobehavioral assessments (MASC, BRIEF, SDQ, Conners 3-SR, and TAP scores) were appraised using Wilcoxon signed rank tests, given that assumptions for parametric testing were violated. P-values were corrected for multiple comparisons within each questionnaire, using the False Discovery Rate (FDR) corrections (Benjamini and Hochberg 1995).

### 2.4 Functional MRI paradigm: Gradual-onset Continuous Performance paradigm (gradCPT)

Following instructions and a short practice session, participants were installed inside the MRI scanner, where visual stimuli were back-displayed on a white screen via a 45° angled mirror fixed to the head coil. Both stimuli presentation and response recording (through MRI-compatible response buttons) were controlled with Psychophysics Toolbox extensions (https://psychtoolbox.org) implemented in Matlab 2019b (Mathworks Inc., Natick, MA, USA).

In the scanner, participants performed two versions of the gradual-onset continuous performance task (gradCPT), presented in a pseudo-randomized order. Each gradCPT version represented a distinct modality i.e., face or scene (see Figure 1). In the face modality, targets were female faces, while nontargets were represented by male faces. In the scene modality, targets were mountain scenes, while nontargets were city scenes. With a total duration of 6 minutes, each version of the gradCPT encompasses a visual stimulus that gradually transitions between target and nontarget stimuli presented at a constant rate. In each trial (1’200ms duration), the image transition (comprising the succession of 16 images in total) took 800ms, followed by a 400ms pause during which the stimulus was fully coherent. For each version of the gradCPT task, visual stimuli were presented randomly, with target stimuli occurring 10% of the time (60 trials) and nontarget ones 90% of the time (540 trials), without allowing the ongoing stimulus to be repeated in the upcoming trial. Participants were instructed to answer as quickly and accurately as possible to each nontarget stimulus (i.e., male faces or city scenes) by pressing the response button, but to withhold response to target stimuli (i.e., female faces or mountain scenes).

**Figure 1:**
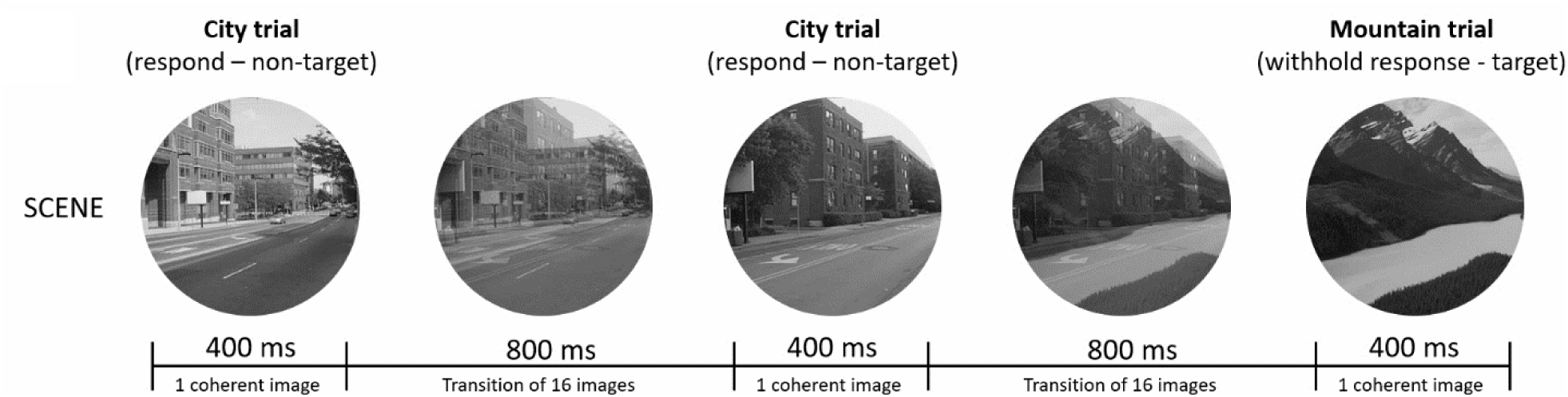
GradCPT. Illustration of the gradCPT paradigm, for the scene task. Target trials (mountain scenes) are presented 10% of the time and need to be withhold, while nontarget trials (city scenes) are presented 90% of the time and need to be answered by pressing a button. For each trial (1’200ms duration), stimulus gradually transition from the current to the next trial during 800ms (succession of 16 images in total), followed by a 400ms pause during the stimulus was fully coherent.

Based on the previous work of Rosenberg, Noonan et al. (2013), a nontarget trial was considered accurate when participants pressed the response button between 560ms and 1’520ms after the stimulus onset. Additionally, ambiguous presses, i.e., highly deviant reaction times (shorter than 560ms or longer than 1’520ms) or multiple button presses during a single trial, were evaluated using the Rosenberg’s iterative algorithm maximizing correct responses and potentially assigned to adjacent trials. Finally, a target trial was considered accurate when participants withheld their response between 560ms and 1’520ms after the beginning of the image transition (stimulus onset).

### 2.5 GradCPT behavioral data analysis

The adolescents’ sustained attention performance was assessed using the accuracy (AC), the sensitivity scores (d’) as a measure of overall performance, as well as the reaction time variability, defined using the coefficient of variation (RT_CV_) as a measure of intraindividual response variability. AC scores were computed using correct commission (CCs; i.e., success to respond to nontarget trial) and correct omission (COs; i.e., success to inhibit responses during target trials) for each participant, for each group, and for each version of the gradCPT separately. d’ scores, computed by contrasting both stimuli types as *z(hit rate) – z(false alarm rate)* (detailed description of d’ calculation can be found in Rosenberg and colleagues (2016)), were assessed for each participant, for each group, and for each version of the gradCPT separately. RT_CV_ was computed as the standard deviation of reaction times divided by the mean reaction times (Rosenberg, Finn et al. 2015, Fortenbaugh, Rothlein et al. 2018, Urbain, Sato et al. 2019) and was assessed for correct nontarget stimuli, for each participant, for each group, and for each version of the gradCPT separately.

Due to their robustness as indicators of attentional performance, statistical analysis of behavioral performance focused solely on d’ and RT_cv_ scores, using R Software and RStudio. Consequently, the article does not delve into the analysis of AC data its results. Sustained attention performance was analyzed by entering d’ and RT_CV_ scores in separate 2 x 2 mixed-model repeated-measures ANOVAs, with modality (face and scene) as within-subject levels and group (VPT and FT) as a between-subject factor.

Additional analyses were performed to investigate the effect of major covariates of interest on sustained attention performance (d’, and RT_CV_ measures). The selection of the covariates was based on the scores resulting from i) neonatal and demographic characteristics and ii) neurobehavioral assessment showing significant differences between the VPT and the FT groups (see Table 1). The covariates include gestational age, birth weight and socio-economic status (neonatal and demographic factors) as well as MASC and SDQ Emotional problems, Peer problems and Prosocial scores (neurobehavioral assessments). Both d’ and RT_CV_ were entered in different 2 x 2 x 1 mixed-model repeated-measures ANCOVAs, with modality as within-subject level, group as a between-subject factor, plus a covariate as a single factor.

**Table 1.** Neonatal and demographic characteristics (**A**), neurobehavioral assessment and computerized attentional tasks (**B**), as well as gradCPT behavioral performances (**C**), for the very preterm (VPT) and full-term (FT) participants included in the study.

|  |  | VPT (n = 34) | FT (n = 28) | Group comparison |
| --- | --- | --- | --- | --- |
| **(A) Neonatal and demographic characteristics** |  |  |  |  |
| Gestational age (weeks) |  | 201.15 ± 12.25 | 277.58 ± 6.74 | W = 884, **p < 0.001** |
| Birth weight (grams) |  | 1166.47 ± 353.88 | 3390.37 ± 366.07 | W = 918, **p < 0.001** |
| Age at assessment (years) |  | 15.69 ± 1.59 | 15.31 ± 2.09 | W = 405, p = 0.32 |
| Sex (n) |  | 19 F ; 15 M | 13 F ; 15 M | X<sup>2</sup> (1, N=62) = 0.54, p = 0.45 |
| Socio economic status (Largo Scale) |  | 4.89 ± 3.42 | 3.11 ± 1.65 | W = 325, **p = 0.04** |
| General ability index (WISC-IV) |  | 108.32 ± 10.90 | 110.67 ± 10.44 | W = 521, p = 0.36 |
| **(B) Neurobehavioral assessment and Computerized attentional tasks** |  |  |  |  |
| Strengths and Difficulties Questionnaire (SDQ – *Parent-report*)* | Emotional problems | 2.19 ± 2.60 | 1.35 ± 2.10 | W = 278, p = 0.18 |
|  | Peer problems | 2.37 ± 1.74 | 1.08 ± 1.09 | W = 190, p = **0.01** |
|  | Prosocial skills | 8.85 ± 1.51 | 8.08 ± 1.47 | W = 236, p = **0.05** |
| Multidimensional Anxiety Scale for Children 2<sup>nd</sup> Edition (MASC 2-SR – *Self-report*) | Total score | 40.24 ± 14.95 | 29.22 ± 10.33 | W = 238, p = **0.002** |
| Conners 3-SR (*Self-report*) (T scores) | Inattention | 56.93 ± 10.54 | 54.81 ± 10.08 | W = 400, p = 0.63 |
|  | Hyperactivity / Impulsivity | 53.72 ± 9.98 | 57.03 ± 11.48 | W = 514, p = 0.63 |
|  | ADHD Index | 48.25 ± 21.06 | 49.00 ± 17.67 | W = 465, p = 0.63 |
| Behavioral Rating Inventory of Executive Function (BRIEF – *Parent-report*)* (T scores) | Behavioural regulation index (BRI) | 53.85 ± 12.44 | 50.69 ± 12.23 | W = 292, p = 0.53 |
|  | Metacognition Index (MI) | 55.67 ± 12.87 | 52.03 ± 10.76 | W = 299, p = 0.53 |
|  | Global Executive Composite (GEC) | 55.59 ± 13.39 | 51.88 ± 10.76 | W = 309, p = 0.53 |
| Test of Attentional Performance (TAP) (T scores) | Go/NoGo – 1 target / 2 stimuli | 41.48 ± 6.45 | 42.48 ± 7.61 | W = 314, p = 0.67 |
|  | Go/NoGo – 2 targets / 5 stimuli | 45.62 ± 9.02 | 46.72 ± 8.23 | W = 332, p = 0.93 |
| **(C) gradCPT behavioral performances** |  |  |  |  |
| Sensitivity scores (d')** | gradCPT – Face | 0.47 ± 0.33 | 0.44 ± 0.42 | t (45.98) = 0.28, p = 0.77 |
|  | gradCPT – Scene | 1.10 ± 0.46 | 1.12 ± 0.59 | t (46.41) = -0.15, p = 0.87 |
| Reaction time Variability (RT<sub>CV</sub>)** | gradCPT – Face | 0.29 ± 0.04 | 0.29 ± 0.05 | t (48.33) = -0.02, p = 0.98 |
|  | gradCPT – Scene | 0.26 ± 0.03 | 0.25 ± 0.04 | t (50.34) = 1.33, p = 0.18 |

Notes: Mean group values are presented with the standard deviation. Regarding neurobehavioral assessment, p-values are resulting from Wilcoxon signed rank test and corrected for multiple comparisons within a single questionnaire, using the False Discovery Rate (FDR). Abbreviation: n = number of participants ; F = female ; M = male ; t = Student t-test ; X<sup>2</sup> = Chi-squared test ; W = Wilcoxon signed rank test ; WISC-V = Wechsler Intelligence Scale for Children-4th Edition. Stars: \* indicate that parent-reported questionnaires (i.e., SDQ and BRIEF) were missing for 6 VPT and 1 FT adolescents ; \*\* indicate that 2 VPT and 2 FT adolescents were excluded from the behavioral analyses, as they presented incoherent results or button responses issues in one of the two gradCPT tasks.

### 2.6 Functional MRI acquisition

Magnetic Resonance Imaging (MRI) acquisition was performed at the Campus Biotech in Geneva (Switzerland) in a 3T MRI scanner (Siemens Magnetom Prisma, Germany) with a standard 64-channel head coil. Functional images were acquired using multi-slice echo-planar imaging (EPI) sequences in single short (*T* ^∗^-weighted; TR/TE = 720/33.0 ms, flip angle = 52°, Voxel dimensions = 2 mm isotropic, FOV = 208 × 208 mm), while a magnetization-prepared rapid acquisition gradient-echo (MP-RAGE) sequence was used for structural images (*T*_1_-weighted; TR/TE = 2300/2.32 ms, flip angle = 8°, Voxel dimensions = 0.90 mm isotropic, FOV = 240 × 240mm). Additionally, a fieldmap was acquired for each participant (TR/TE_1_/TE_2_ = 627/5.19/7.65ms, flip angle = 60°, Voxel dimensions = 2mm isotropic, FOV = 208 x 208 mm).

### 2.7 Functional MRI data preprocessing

Preprocessing and statistical analyses of MRI images were performed using Statistical Parametric Mapping 12 tool (SPM12, Wellcome Trust Centre for Imaging, London, UK) implemented in Matlab R2021b (The MathWorks, Inc., Natick, Massachussets, USA). Following a conversion from native DICOM to NIFTI format, fMRI images were preprocessed via an in-house pipeline (previously described in (Liverani, Freitas et al. 2020, Freitas, Liverani et al. 2021). Briefly, fMRI images were spatially realigned, unwarped to correct for potential geometric distortions allowing to improve both co-registration and spatial normalization (Hutton, Bork et al. 2002), and co-registered to their corresponding structural images. Structural images were then segmented with the SPM12 segmentation algorithm to identify distinct tissue probability maps, i.e., grey matter, whiter matter and cerebrospinal fluid (Ashburner and Friston 2005) used to generate a study-specific template (using Diffeomorphic Anatomical Registration using Exponential Lie algebra) (DARTEL; Ashburner 2007). Finally, functional images were normalized to the DARTEL template and smoothed with a 6-mm isotropic Gaussian kernel. Additionally, the extent of head motion from volume to volume was computed using the mean framewise displacement (FD; Power, Barnes et al. 2012, Power, Mitra et al. 2014), for each experimental modality of the gradCPT. Participants with more than 20% of the frames affected by motion, i.e., frames with a mean FD above 0.5mm as well as the one preceding and one subsequent frame, were removed from further analyses. Based on this motion criterion, 1 FT control participant was excluded from the scene gradCPT.

### 2.8 Functional MRI data analysis

For each experimental modality separately, the first-level analysis was performed using a general linear model (GLM) to model all trials from the two experimental stimuli, i.e., target (T) and nontarget (NT). Trial onset was aligned on the beginning of the image transition, with a duration 0.57sec, and convolved with the canonical hemodynamic response function (cHRF). Motion parameters, as estimated during the realignment pre-processing step (six realignment values) were entered as non-interest covariates. For each participant and each experimental modality, T-contrast of interest was computed by comparing the target condition with the nontarget condition (T > NT) at the first level.

The second-level whole-brain analysis was performed for each experimental modality separately using a flexible factorial design with stimulus (T and NT trials), group (VPT and FT) and subject as separate factors. Sustained attention was assessed via response inhibition processes. The main effect of response inhibition was determined by comparing target to nontarget conditions (T > NT). First, the cerebral network associated with response inhibition and shared by the two groups (i.e., similarities) was assessed with a conjunction analysis (VPT (T > NT) & FT (T > NT)). Second, the effect of prematurity on the response inhibitory network was assessed by comparing the cerebral activation between the two groups (VPT (T > NT) > FT (T > NT) or FT (T > NT) > VPT (T > NT)). Inclusive masks were used to confirm the direction of the interaction.

Then, we explored how cerebral activations within the response inhibition network varied as a function of covariates of interest by performing between-group univariate linear regression analyses. As described for behavioral analyses, the selection of covariates was based on scores resulting from i) neonatal and demographic characteristics (gestational age, birth weight and socio-economic status) and ii) significant differences between the VPT and the FT groups in neurobehavioral measures (MASC and SDQ scores) (see Table 1). All values were entered as parametric covariates in separate SPM analyses. Finally, the direction of significant peak activations resulting from the univariate linear regression analyses between the response inhibition network and covariates of interest was confirmed by additional post-hoc analyses. For each participant, beta values, reflecting the main effect of response inhibition (T > NT), were extracted using spheres (27 voxels) centered on the main peak coordinates identified by covariates analyses in SPM. Correlations analyses between the resulting beta values and corresponding covariates of interest were then performed for each group (VPT or FT) separately, using R Software and R Studio.

For all second-level whole-brain analyses, we report brain activations with uncorrected p-values thresholded at p < .001, with a cluster size of > 50 continuous voxels (Lieberman and Cunningham 2009).

## 3. Results

### 3.1 Participants

Of the 71 enrolled participants, 8 participants (VPT = 7; FT = 1) were excluded as they did not complete the MRI session entirely, and 1 participant (VPT = 1) was excluded because of issues with the button device in the two versions of the gradCPT. The final sample comprised 62 participants, including 34 VPT and 28 FT participants, aged between 11 and 18 years (Table 1).

### 3.2. Neonatal/General cognition/Demographic characteristics

Age at assessment as well as general intellectual abilities were comparable between the two groups of participants (p > .32 and p > .36, respectively). The proportion of female and male participants was similar between groups (p > .45). Socio-economic status, measured using the Largo score (Largo et al., 1989), revealed lower socio-economic status (higher Largo scores) in the VPT compared to the FT group (p = .04). Neonatal/Demographic characteristics scores and p-values between the two groups are presented in Table 1A.

### 3.3 Neurobehavioral assessment

Statistical analyses revealed significant group differences in behavioral and socio-emotional outcomes measured using the SDQ parent-reported questionnaire, with higher scores in the VPT compared to the FT adolescents, regarding Peer problems (p = .01) and Prosocial scale (p = .05). No significant difference was observed between the VPT and FT groups regarding SDQ-Emotional problems (p > .05). Global assessment of anxiety using the MASC showed significantly higher scores in the VPT compared to the FT adolescents (p = .002). Executive and attentional abilities measured using the BRIEF, the Conners 3-SR and the TAP were comparable between the two groups across all scores (p > .05). Outcomes of the neurobehavioral assessment are presented in Table 1B.

### 3.4 GradCPT behavioral performance

Accuracy on gradCPT performance across both groups revealed a mean correct commission (i.e., correct press during non-target condition) of 88.44% (SD = 13.64%) (face: M = 83.71%; SD = 16.18%; scene: M = 93.17; SD = 8.26%), as well as a mean correct omission (i.e., correct withhold during target condition) of 28.24% (SD = 21.03%) (face: M = 26.19%; SD = 19.74%; scene: M = 30.27%; SD = 22.22%), for both groups combined. Statistical analysis of the gradCPT performance revealed a significant main effect of modality, showing higher d’ scores [F_(1, 56)_ = 80.74; p < .001] and lower reaction time variability [F_(1, 56)_ = 54.26; p < .001] for the scene gradCPT (d’: M = 1.11; SD = 0.51 ; RT_CV_: M = 0.25; SD = 0.03) compared to the face gradCPT (d’: M = 0.46; SD = 0.37 ; RT_CV_: M = 0.29; SD = 0.04). Both groups showed comparable d’ and RT_CV_ scores (p > .5). Behavioral data revealed no group by modality interaction (p > .2). Sustained attention performance, i.e., d’, and RT_CV_ scores is presented for each gradCPT modality and for each group separately, in Table 1C.

Covariate analyses on adolescents sustained attention performance (d’, and RT_CV_ scores) revealed no significant effect of prematurity (group by covariate interaction) associated with i) neonatal and demographic characteristics (gestational age, weight at birth, socio-economic status), nor ii) neurobehavioral assessment (MASC and SDQ scores) (p > .5).

### 3.5 fMRI results / Whole-brain activations during the gradCPT paradigm

#### 3.5.1 Between-group similarities and differences in activations related to Response Inhibition

In the face gradCPT (Table 2A, Figure 2A), the main effect of response inhibition (target > nontarget) shared by both groups (VPT & FT) revealed common right-lateralized prefrontal activations in the dorsal premotor cortex (dPMC) and the supplementary motor area (SMA), as well as in the bilateral anterior midcingulate cortices (aMCC). The direct between-group comparison showed higher activation for the VPT compared to the FT in the right occipital face area (OFA). No decrease of cerebral activity was found in the VPT compared to the FT group, regarding the face modality.

**Figure 2:**
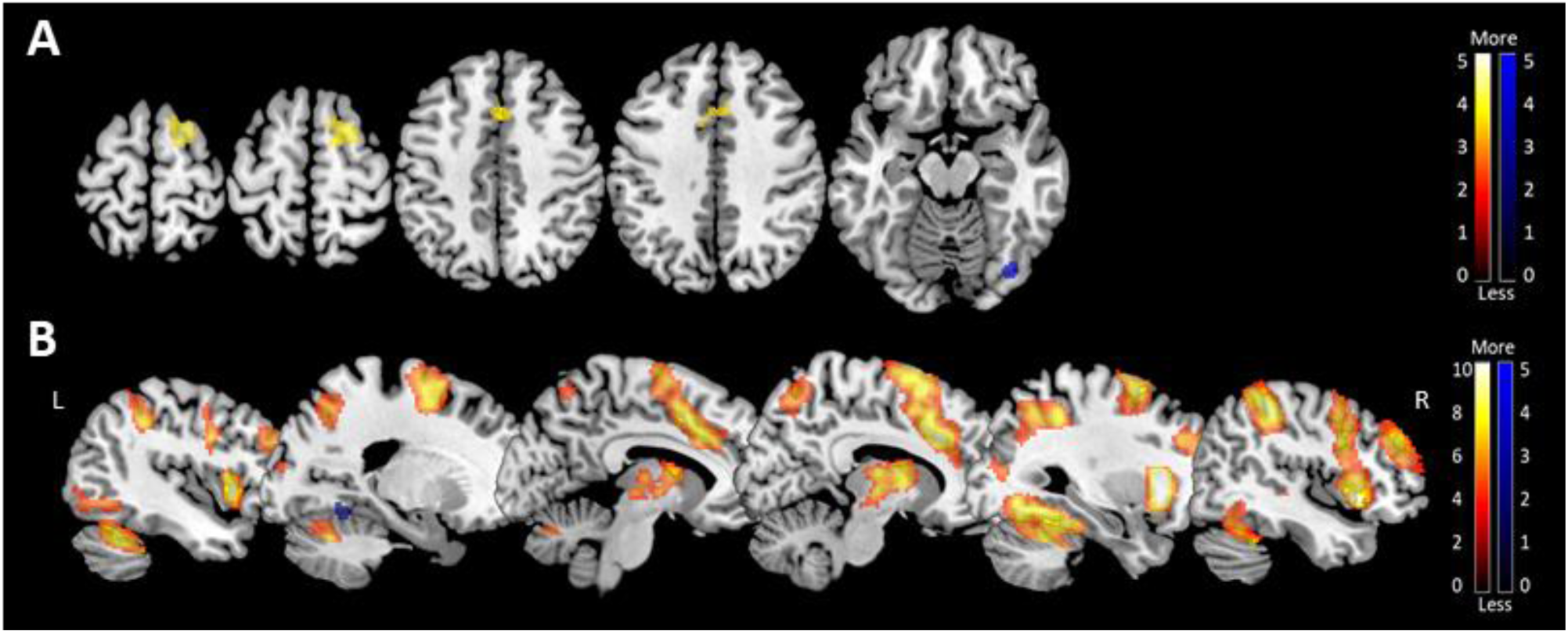
Brain activation engaged by sustained attention during response inhibition trials (target > nontarget) in the gradCPT paradigm is presented as the degree of brain activation, for Face task (A) and Scene task (B) separately. Cerebral areas commonly activated across very preterm (VPT) and full-term (FT) adolescents are presented in a red-yellow gradient, while greater increases observed in the VPT compared to the FT group in the right-Fusiform gyrus (**A** ; Face task) and in the left-Lingual gyrus (**B** ; Scene task) is depicted in blue. All findings are presented in MNI space. All clusters are significant at the peak-level at p < .001, uncorrected for multiple comparisons, with a minimum size of 50 voxels.

**Table 2.** Localization (MNI coordinates) and peak activation values (z score) for brain areas engaged during the GradCPT paradigm and separately analyzed for the Face (A) and the Scene (B) tasks. For each task, shared activations across both groups (VPT & FT) are reported in the upper part of the table, while differential activations (between-group analyses) are reported in the bottom part. Brain activations are calculated for the main effect of response inhibition (target > nontarget).

| Region |  | Lat. | Z score | MNI Coordinates |  |  |
| --- | --- | --- | --- | --- | --- | --- |
|  |  |  |  | x | y | z |
| **(A) FACE Task – Response Inhibition (target > nontarget)** |  |  |  |  |  |  |
| **Common activations across both groups** |  |  |  |  |  |  |
| Frontal | Superior – Dorsal Premotor cortex | R | 4,05 | 22 | 8 | 68 |
|  | Medial – Supplementary motor area | R | 3,79 | 12 | 0 | 58 |
|  | Medial – anterior Midcingulate cortex | R | 3,69 | 6 | 18 | 42 |
|  | Medial – anterior Midcingulate cortex | L | 3,58 | -8 | 12 | 38 |
| **Very Preterm > Full Term** |  |  |  |  |  |  |
| Occipital-Temporal | Lateral – Occipital face area | R | 4,37 | 36 | -76 | -16 |
| **(B) SCENE Task – Response Inhibition (target > nontarget)** |  |  |  |  |  |  |
| **Common activations across both groups** |  |  |  |  |  |  |
| Frontal | Medial – Supplementary motor area | R | 6,20 | 14 | 2 | 68 |
|  | Medial – anterior Midcingulate cortex | R | 6,60 | 2 | 14 | 46 |
|  | Medial – anterior Midcingulate cortex | R | 6,88 | 8 | 22 | 32 |
|  | Middle - Dorsolateral prefrontal | R | 5,78 | 38 | 44 | 24 |
|  | Middle - Dorsolateral prefrontal | L | 4,87 | -38 | 34 | 30 |
|  | Inferior - Pars opercularis | R | 6,45 | 52 | 10 | 16 |
| Parietal | Superior - Precuneus | R | 4,49 | 10 | -62 | 58 |
|  | Superior - Precuneus | L | 4,97 | -22 | -62 | 44 |
|  | Inferior – Supramarginal gyrus | R | 6,77 | 48 | -34 | 48 |
|  | Inferior – Supramarginal gyrus | L | 5,10 | -48 | -36 | 44 |
| Temporo-Parietal Junction | Inferior parietal gyrus | R | 5,28 | 28 | -58 | 46 |
|  | Inferior parietal gyrus – Angular gyrus | L | 5,28 | -30 | -54 | 54 |
| Temporal | Inferior gyrus | R | 4,78 | 54 | -52 | -8 |
| Occipito-Temporal | Lateral – Fusiform gyrus | R | 6,45 | 34 | -42 | -22 |
|  | Lateral – Fusiform gyrus | L | 6,51 | -30 | -46 | -16 |
| Occipital | Middle gyrus | L | 4,95 | -32 | -80 | 12 |
| Other | Posterior – Cerebellum (VI) | R | 5,98 | 36 | -50 | -30 |
|  | Posterior – Cerebellum (VI) | L | 6,87 | -36 | -54 | -28 |
|  | Anterior Insula | R | 7,44 | 40 | 16 | -4 |
|  | Anterior Insula | L | 6,98 | -38 | 12 | -2 |
| **Very Preterm > Full Term** |  |  |  |  |  |  |
| Occipital-Temporal | Medial – Lingual gyrus | L | 4,50 | -20 | -54 | -12 |

Notes: All reported peaks are significant at $p < .001$ uncorrected for multiple comparisons with cluster extent > 50 voxels. Abbreviation: Lat.: Hemisphere lateralisation ; Unc.: Uncorrected. Z-score values refer to the activation maxima to the SPM coordinates. 3 participants (VPT = 1; FT = 2) were excluded from the fMRI face or scene gradCPT task analysis as they presented incoherent results at the behavioral level or high level of motion artefacts ( $FD > 20\%$ ).

In the scene gradCPT (Table 2B, Figure 2B), the main effect of response inhibition shared by both groups revealed a widespread fronto-parieto-occipital brain network. This network of activations encompassed predominantly right-lateralized activations located in medial frontal areas, including both SMA and aMCC, the dorsolateral prefrontal cortices (DLPFC), the inferior frontal gyrus (IFG) as well as bilateral parietal activations in the superior and the inferior parietal gyri (SPG and IPG, respectively). In addition, the middle occipital gyrus (MOG) was activated in the left hemisphere, while bilateral activations were found in the temporo-parietal junction (TPJ) and in the occipito-temporal junction (i.e., fusiform gyri), as well as in the posterior cerebellum and in the anterior insula. The direct between-group comparison showed higher activation for the VPT compared to the FT in the left lingual gyrus. No decrease of cerebral activity was found in the VPT compared to the FT group, regarding the scene modality.

#### 3.5.2. Relationship between brain activation related to Response Inhibition and neonatal/demographic characteristics and neurobehavioral abilities

Linear parametric regression analyses were performed to investigate how individual differences in neonatal/demographic characteristics and neurobehavioral assessments were related to distinct cerebral activity patterns (target > nontarget) between VPT and FT adolescents.

In the face gradCPT, regression analyses revealed a stronger positive association between MASC scores (higher anxiety scores) and right-lateralized posterior activations, i.e., lingual gyrus, insula and posterior cerebellum, as well as in the left MOG in the VPT compared to the FT group (Table 3A, Figure 3A). Additional beta-extraction indicated between-group opposite associations in the right-lateralized lingual gyrus and in the cerebellum, showing positive correlations in the VPT and negative correlations in the FT adolescents (Table 3A, Figure 3A). Finally, the analysis of the SDQ subscales revealed higher positive associations between Peer problems scores, indicating larger relationship difficulties with peers, and a widespread fronto-occipito-cerebellar brain network, including right-lateralized SMA, frontopolar cortex, middle frontal gyrus (MFG) and the left IFG, in the FT compared to the VPT group (Table 3B, Figure 3B). Beta-extraction analyses revealed that the activation in the right MFG, as well as the left-posterior cerebellum (VI and Crus1) were negatively correlated with the Peer problem scores in VPT, but positively correlated in the FT group (Table 3B, Figure 3B).

**Figure 3:**
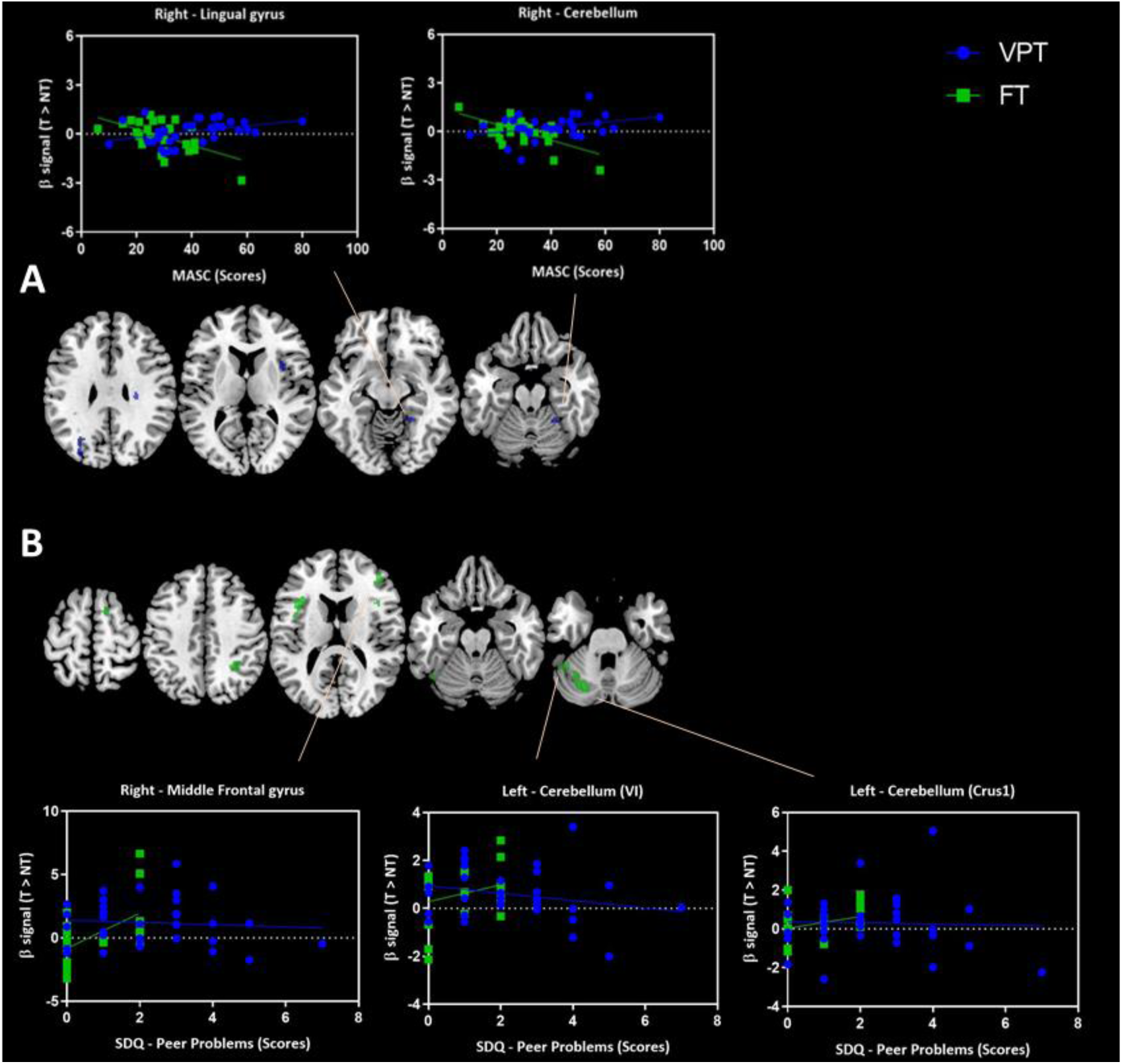
Illustrations of the cerebral areas activated by sustained attention during response inhibition trials (target > nontarget) in the gradCPT Face task, and whose activity showed positive correlations with neurobehavioral assessments and gradCPT behavioral performances in whole-brain SPM analyses and presented as a degree of brain activation. Greater positive associations between cerebral recruitments and (**A**) higher (worse) anxiety scores using the Multidimensional Anxiety Scale (MASC) (in blue) and (**B**) higher (worse) peer problems scores using the Strength and Difficulties Questionnaire (SDQ) (in green). For each activation peak presenting between-group opposite patterns of correlation, individual neurobehavioral scores as a function of beta signal associated with response inhibition (target > nontarget) are plotted, i.e., right Lingual gyrus and right Cerebellum for MASC scores (**A**) as well as right Middle frontal gyrus and left Cerebellum for SDQ-Peer problems scores (**B**). VPT outcomes are depicted in blue, while FT outcomes are in green. All findings are presented in MNI space. All clusters are significant at the peak-level at p < .001, uncorrected for multiple comparisons, with a minimum size of 50 voxels.

**Table 3.**
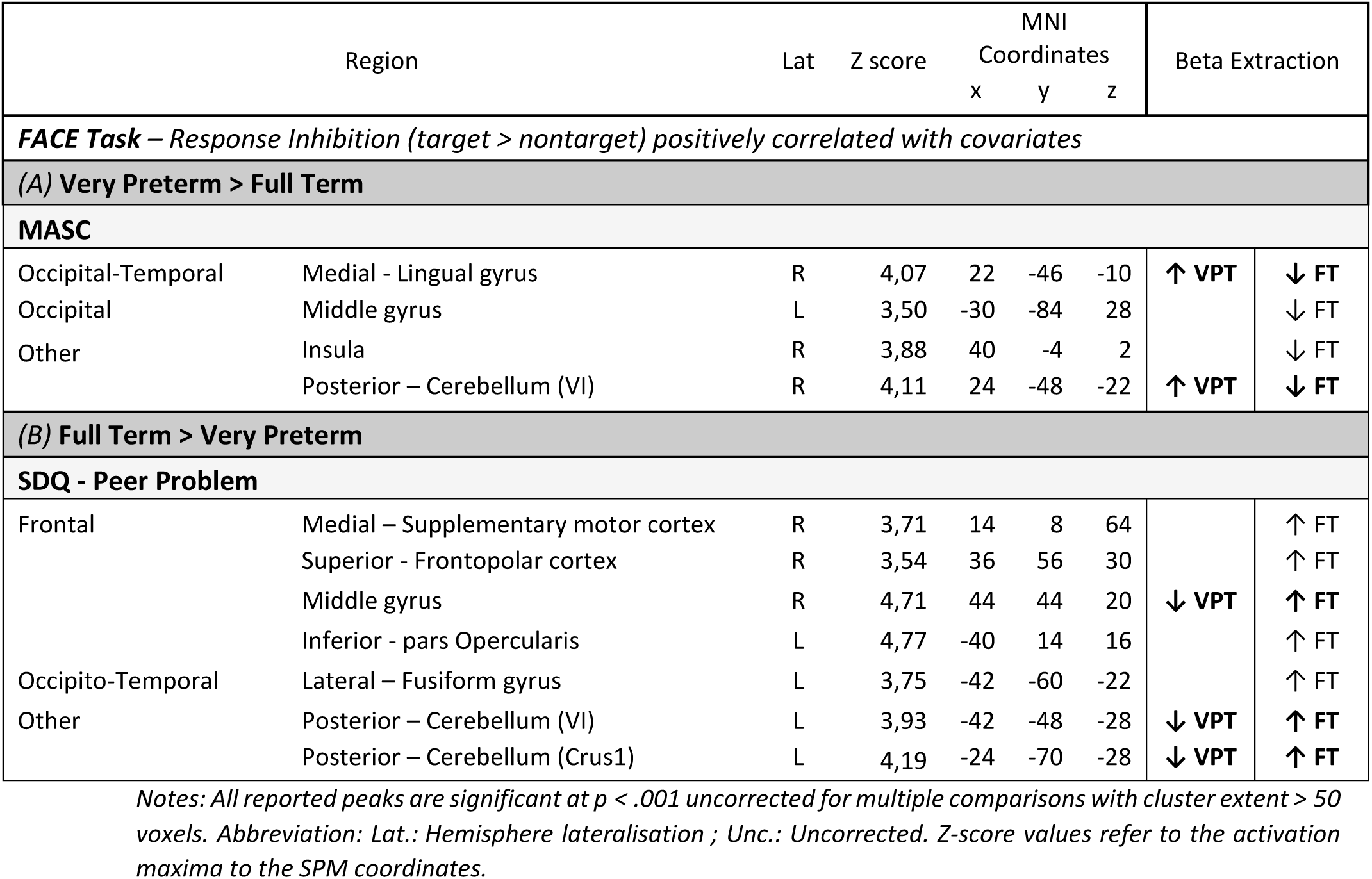
Localization (MNI coordinates) and peak activation values (z score) for brain areas engaged during response inhibition trials (T > NT) in the gradCPT Face task and whose activity showed a positive correlation with neurobehavioral assessments and gradCPT behavioral performances in whole-brain SPM analysis. (**A**) Greater positive associations between cerebral recruitments and higher (worse) anxiety scores using the Multidimensional Anxiety Scale (MASC) or higher variability in reaction times (RTcv; i.e., lower gradCPT attentional performances) in the very preterm (VPT) compared to the full-term (FT) participants. (**B**) Greater positive associations between cerebral recruitments and higher birth weight (beneficial) or higher (worse) peer problems scores using the Strength and Difficulties Questionnaire (SDQ) in the FT compared to the VPT participants. For each peak activation, the direction of the association (i.e., significant correlations between brain areas and covariates of interest), assessed using beta-extraction analyses and correlations analyses in each group separately, is reported in the right part of the table. Arrow pointing up for positive correlations and pointing down for negative correlations. Peak activations for which the direction of the association show opposite results between the two groups are presented in bold.

Regression analyses on the scene gradCPT revealed no distinct between-group associations between cerebral activations and covariates of interest.

## 4. Discussion

In the present study, we examined how prematurity impacts cerebral activity linked to visual sustained attention and its relationship with attentional and socio-emotional competences, which are known to be affected in VPT individuals. To this aim, VPT and FT adolescents completed two distinct version of the gradCPT, involving face and scene processing, enabling the exploration of sustained attention components through response inhibition processes. Overall, our results demonstrated preserved processing of sustained attention in VPT adolescents compared to their age-matched FT peers, as evidenced by comparable behavioral performance and task-related brain activations. Nevertheless, further investigations uncovered heightened cerebral activations in posterior sensory areas among VPT compared to their FT counterparts. Moreover, in VPT adolescents, difficulties in emotion regulation and increased peer relationship problems were associated with atypical recruitment of fronto-occipito-cerebellar regions.

### Neurobehavioral and attentional assessment

Compared to FT controls, VPT adolescents in our cohort showed difficulties in socio-emotional functioning in their daily lives, including higher incidences of peer problems and elevated anxiety levels, as measured through both parent-reported SDQ and self-reported MASC questionnaires. These findings are consistent with previous research (Sømhovd, Hansen et al. 2012, Johns, Lacadie et al. 2019, Urbain, Sato et al. 2019, Schnider, Disselhoff et al. 2020, Siffredi, Liverani et al. 2023a, Bilgin, Wolke et al. 2024). Executive and attentional competences (i.e., BRIEF-parent, Conners 3-SR and TAP) were similar between our two groups , corroborating some research (Sølsnes, Skranes et al. 2014), but contradicting others (Burnett, Anderson et al. 2018, Doyle, Spittle et al. 2021). Remarkably, the difficulties identified seem to be confined to socio-emotional competences in the current cohort of VPT adolescents. This observation is noteworthy, particularly considering that adolescence represents a pivotal developmental stage characterized by enhanced sensitivity to social evaluation and peer influence (Somerville 2013). Previous studies have frequently reported difficulties in socialization and socio-emotional competences among preterm adolescents (Schmidt, Miskovic et al. 2008, Montagna and Nosarti 2016), which are consistent with our findings.

Regarding the gradCPT experimental task, we found better behavioral performance in the scene compared to the face modality, across both groups. This was evidenced by higher sensitivity (d’) and lower reaction times variability (RT_CV_). These results are particularly noteworthy, as to our knowledge, this is the first direct comparison of the performance scores between these two gradCPT modalities. Our findings imply that the scene gradCPT requires less cognitive effort than the face gradCPT. Various interpretations could elucidate this observation. First, this result might be attributed to a more evident distinction between the two types of stimuli in the scene modality(i.e., cities and mountains) than the ones in the face modality (i.e., male and female faces), triggering exogenous attention which might in turn facilitates the discrimination of the scene stimuli . Second, our adolescents might have encountered greater difficulties in allocating attentional resources to local visual elements of faces required for gender discrimination compared to the more global features inherent in cities or mountains images. This difficulty may stem from a visual attention bias towards global processing, a phenomenon documented from late childhood to adulthood in prior research (Poirel, Simon et al. 2011). Third, the diminished performance in the current face version of the gradCPT might be attributed to the relatively delayed maturation of configural face processing, which is crucial for facial identity recognition and necessitates the complex integration of facial features (Mondloch, Le Grand et al. 2002, Fuhrmann, Knoll et al. 2016).

Critically, attentional scores were comparable between the VPT and the FT groups, suggesting no adverse impact of prematurity on sustained attention performance in the gradCPT. Although attentional difficulties are frequently reported in the VPT population, our current findings corroborate with those of other researchers who have also observed preserved attentional behavioral performance (Daamen, Bäuml et al. 2015), included in response inhibition processes in VPT children (Urbain, Sato et al. 2019), adolescents (Nosarti, Rubia et al. 2006, Lawrence, Rubia et al. 2009) and young adults (Olsen, Dennis et al. 2018, Réveillon, Hüppi et al. 2018).

Interestingly, we found no significant correlation between behavioral attentional performance (i.e., RTcv and d’) resulting from the two versions of the gradCTP, and the covariates of interests. These covariates encompassed neonatal characteristics inherent to prematurity (such as gestational age and birth weight), socio-economic status, and socio-emotional difficulties (as measured by MASC and SDQ scores). Altogether, these results confirm preserved sustained attention performance in our cohort of VPT adolescents at the behavioral level, thus supporting conclusions drawn from parent-reported questionnaires assessing attention and executive functions.

### Between-group similarities in cerebral activation during response inhibition

Examining brain activity during the face gradCPT, we observed activation within a frontal network, including the right dPMC, SMA, as well as the bilateral MCC, activations in both VPT and FT adolescents during the response inhibition of target stimuli. Previous studies have emphasized the importance of various brain areas, including the MCC, in continuously maintaining attentional focus (Breckel, Giessing et al. 2011, Langner and Eickhoff 2013). By showing comparable recruitment of the MCC area between our two groups, the present study suggests that VPT adolescents were able to sustain their focus of attention over time to the same extent as the controls. Additionally, activations in dPMC and SMA areas indicate that both VPT and FT adolescents were capable of enhancing goal-directed preparation and motor control, necessary for inhibiting external target stimuli. These activations are in line with previous work using visual paradigms requiring the withholding of motor response, as the present experimental task (Schubotz and von Cramon 2001, Nachev, Kennard et al. 2008, Nakayama, Sugawara et al. 2022).

Regarding the scene gradCPT, both VPT and FT participants similarly activated an extended and mostly bilateral fronto-parieto-occipital network frequently engaged in several paradigms assessing sustained attention in neurotypical adults (Langner and Eickhoff 2013) and young adolescents (Morandini, Silk et al. 2020). Previously reported in alerting processes, areas in the frontoparietal network, in particular DLPFC, MCC, IFG, and TPJ, are essential to allocate and maintain attentional resources to unexpected but relevant targets (Fan, McCandliss et al. 2005, Langner and Eickhoff 2013, Morandini, Silk et al. 2020). Our participants also recruited a parieto-occipital network, encompassing the SPG and fusiform gyri, which have been described in previous fMRI studies during the allocation of visual attention to sensory information (Fan, McCandliss et al. 2005). Interestingly, the response inhibition contrast in the scene gradCPT engaged the anterior part of the insular cortex, which is a component of the salience network (also known as the cingulo-operculum network), recognized as crucial for detecting task-relevant stimuli and engaging required processes (Namkung, Kim et al. 2017). In conjunction with behavioral outcomes, the involvement of the anterior insula in the scene gradCPT but not in the face gradCPT, lends support to our hypothesis. Namely, the distinction between target and nontarget visual stimuli in the scene gradCPT (i.e., cities vs. mountains) appears to evoke heightened salience compared to the face gradCPT (i.e., male vs. female faces), potentially enhancing stimulus discrimination. This observation remains consistent regardless of adolescents’ premature birth status.

It is noteworthy that sustained attention processes are frequently characterized by predominantly right-hemisphere cerebral activation (Langner and Eickhoff 2013, Morandini, Silk et al. 2020). However, a study examining vigilance processes, a term often used interchangeably with sustained attention, also emphasized that this right cerebral lateralization might result from employing relatively simple attentional tasks (Helton, Warm et al. 2010). Specifically, the authors demonstrated that increasing task difficulty (for instance, by reducing the salience of the targets and thus their discriminability) would diminish unilateral recruitment and induce a shift to more symmetrical patterns of cerebral activity. Hence, both bilateral cerebral activation and overall low accuracy scores in target conditions observed in this study converge to indicate that the current gradCPT paradigm presents a challenging attentional task. Recruiting both hemispheres may provide additional processing power to effectively accomplish the task.

Overall, behavioral and neuroimaging outcomes stemming from the face and the scene versions of the gradCPT demonstrated preserved sustained attention in VPT adolescents. This was evidenced by the engagement of task-related cerebral areas, typically involved when performing of sustained attention tasks, and similarly activated in both groups.

### Between-group differences in cerebral activation during response inhibition

Contrasting with previous evidence highlighting reduced activations in VPT adolescents (Nosarti, Rubia et al. 2006, Griffiths, Gundersen et al. 2013), the current study showed no significant cerebral decrease during response inhibition in our VPT population compared to the full-terms (Daamen, Bäuml et al. 2015, Olsen, Dennis et al. 2018). However, the direct between-group comparison indicated greater brain areas recruitment in the VPT compared to FT adolescents. More particularly, the right OFA and left lingual gyrus were more activated when performing the face and the scene gradCPT, respectively. The OFA is usually engaged during the perception of facial components (e.g., eyes or mouth) (Pitcher, Walsh et al. 2011), while the lingual gyrus is implicated in the detection of simple features of visual stimuli (e.g., contours) (Felleman and Van Essen 1991). Increased activations in these extrastriate occipital regions might reflect greater target stimuli processing in sensory areas (Hopfinger, Buonocore et al. 2000). The posterior hyper-activation pattern found here aligns well with other works on response inhibition reporting larger activations in temporo-occipital areas in adolescents (Nosarti, Rubia et al. 2006) and young adults born prematurely (Lawrence, Rubia et al. 2009, Olsen, Dennis et al. 2018). Similar associations were also found between enhanced lateral occipital recruitments and lower gestational age in VPT young adults (Daamen, Bäuml et al. 2015, Olsen, Dennis et al. 2018). Furthermore, prior evidence on a neurotypical population also reported that children rely more on posterior brain regions compared to adults during alerting conditions (Konrad, Neufang et al. 2005). Additionally, typically developing children display more diffuse neural activations compared to adults, who tend to rely more on task-relevant fronto-parietal regions (Durston and Casey 2006). Hence, the observed increase of activation in posterior visual regions can be interpreted as compensatory mechanisms in VPT adolescents, involving enhanced bottom-up visuo-spatial processing, presumably due to an immature network (Nosarti, Rubia et al. 2006, Réveillon, Hüppi et al. 2018, Morandini, Silk et al. 2020).

### Association between cerebral activation during response inhibition and prematurity-related characteristics and difficulties

In this study, we further examined how the pattern of brain activity engaged during a sustained attention task might vary based on the severity of prematurity characteristics and associated difficulties, particularly socio-emotional weaknesses. Although no associations were observed between attentional performance (measured by d’ and RT_CV_) and the covariates of interest (such as prematurity characteristics and difficulties) at the behavioral level, significant associations emerged between these covariates and cerebral activations. fMRI analyses unveiled significant between-group correlations, indicating subtle changes in neuronal activation in VPT adolescents when considering their socio-emotional weaknesses, compared to their FT peers.

More particularly, VPT adolescents with higher anxiety scores (as assessed by the MASC questionnaire) tended to exhibit over-recruitment of right-lateralized medial occipital areas and the cerebellum during response inhibition in the face gradCPT, while FT adolescents displayed opposite negative correlations. This is consistent with similar associations reported between posterior hyper-activations and higher anxiety scores in VPT young adults during a CPT, although this effect was restricted to the precuneus and posterior cingulate cortex (Olsen, Dennis et al. 2018). Paradoxically, VPT individuals presenting peer relationship problems (as indicated by higher peer problems scores in the SDQ scale) showed stronger deactivations of the right MFG and the left posterior cerebellum areas during response inhibition in the same face gradCPT. The involvement of the right MFG during the face gradCPT is of great interest as this region has been identified as critical in sustained attention processing (Song, Lin et al. 2019) as well as in emotion regulation (Blair, Smith et al. 2007) in healthy populations. However, we observed here stronger MFG deactivations in the VPT adolescents presenting higher peer relationship problems. This result is closely related to prior research also reporting the MFG to be deactivated in VPT compared to FT children during successful response inhibition control. These findings suggest that VPT participants, particularly those with higher socio-emotional weaknesses, might present subtle alterations in the MFG activation during sustained attention processes. Additionally, there is evidence of the cerebellum’s involvement in social information processing (Montagna and Nosarti 2016), affective processing (Pierce, Thomasson et al. 2023), and attentional processing (Gottwald, Mihajlovic et al. 2003). However, the reason why the cerebellum exhibited opposite cerebral associations with anxiety scores (i.e., hyperactivation in the right hemisphere) compared to poor social skills (i.e., hypoactivation in the left hemisphere) in presence of social information (i.e., face gradCPT) in VPT adolescents remains unclear and warrants further investigations.

Our observations regarding the impact of prematurity-related difficulties on sustained attention imply that, although behavioral performance remained preserved in VPT adolescents with heightened socio-emotional vulnerabilities (as evidenced by no correlation between behavioral attentional performance and anxiety or peer problem scores), discernible alternations exist in cerebral activity among VPT individuals. Specifically, the increased occipito-cerebellar engagement related to higher anxiety might reflect greater demands to maintain the focus of attention over time. Conversely, reduced fronto-cerebellar engagement related to diminished socio-emotional skills might be indicative of an inability to engage specific systems at an optimal level during the face gradCPT, likely due to an immature network. Nevertheless, this pattern of activity engaged during sustained attention appears to be specific to the socio-emotional context, as no differential association was found between the VPT and their FT peers in the discrimination task presenting no social information (i.e., the scene gradCPT). This specific finding during the face gradCPT is in accordance with previous evidence linking atypical socio-emotional development to impairments in attention orienting abilities in the VPT population (Montagna and Nosarti 2016).

## 5. Limitations

While shedding new light on the cerebral engagement underlying sustained attention in VPT young adults, the present study is subject to limitations that need to be considered. Notably, the reported behavioral accuracy in the target condition are relatively low across all participants. Consistent with previous studies employing CPT paradigms in young adults (Esterman, Noonan et al. 2013, Rosenberg, Noonan et al. 2013), we observed a higher incidence of commission errors (CEs; i.e., failure to inhibit responses during target trials) than omission errors (OEs; i.e., failure to respond to nontarget trial) in both scene and face versions of the gradCPT. However, compared to previous works on gradCPT (M = 20-26%; range = 4% - 49%) (Esterman, Noonan et al. 2013, Rosenberg, Noonan et al. 2013) the number of CEs observed in the current study was surprisingly high overall (face: M = 72%; SD = 21%; scene: M = 68%; SD = 22%), particularly in the face modality of the gradCPT, leading to low d’ scores (d’ < 1). While these scores might be attributable to chance performance or disinterest toward the task, we refute this hypothesis for several reasons. Firstly, participants were unaware of the substantial discrepancy in trial occurrence between conditions, where 10% involved targets and 90% involved nontargets, for both the face and scene gradCPT. Answering randomly would have led to an error rate near 50% in the nontarget condition (OEs), yet this was not observed (< 18%), suggesting a systematic response pattern rather than random responding. Secondly, participants reported feeling more confidence in successfully performing the scene gradCPT compared to the face gradCPT, suggesting that they encountered more difficulties with the latter. This informal observation was based on their verbal feedback following task completion. Simultaneously, the neuroimaging outcomes observed for this scene gradCPT closely align with the sustained attention network described in the literature, suggesting effective attentional engagement with the visual stimuli as instructed. Consequently, we argue that the high incidence of CEs (i.e., failures in inhibiting target stimuli) in the current scene gradCPT is unlikely to be attributed to a lack of interest in the task. Instead, we posit that these high CEs might reflect motor impulsivity in both groups (Halperin, Wolf et al. 1991, Fortenbaugh, DeGutis et al. 2015), indicated by shorter reaction times found in the present work (M = 550ms), compared to those in previous studies (M = 906ms) (Rosenberg, Noonan et al. 2013). This latter speculation is supported by Fortenbaugh and colleagues (2015) who elucidated diverse strategies employed across the lifespan when engaging in sustained attention tasks. Their investigation spanned 60 years, from 10 to 70 years old, utilizing the same scene gradCPT task employed in our study. Their findings highlighted that adolescents, particularly those aged 14 to 17 years, tend to approach the task with greater speed and less cautiously compared to adults, who adopt a slower and more conservative strategy. This variance in approach likely contribute to the overall low accuracy in the target condition observed here, particularly in the scene gradCPT. For all these reasons, we genuinely believe that our young participants were actively engaged in both versions of the experimental gradCPT, exerting efforts to perform at their best despite the observed challenges and possible age-related change in strategy to complete the task.

## 6. Conclusion

In summary, our study reveals that VPT adolescents demonstrate preserved sustained attention processes, evidenced by comparable attentional performance (as measured by gradPCT, Conners questionnaire, and TAP), and expected task-related neuronal patterns of activation with FT controls during the two versions of the gradCPT, each representing distinct modalities. However, the specific observation of posterior over-recruitment in this study suggests that VPT adolescents may exhibit a less mature attentional network than their FT counterparts. The fact that VPT adolescents in our cohort showed general intellectual and attentional functioning within the average range, and comparable to the FT control group, may explain why our study did not entirely replicate previous work highlighting prematurity-related changed across a wide range of brain areas during response inhibition. Furthermore, the enhanced bottom-up visuo-spatial processing observed in this at-risk population might act as a compensatory adaptation, enabling them to achieve performance levels similar to their FT peers. By linking neurobehavioral weaknesses to the cerebral activations during two sustained attention tasks, the present study unveils subtle alterations in brain activity among VPT adolescents. Specifically, we highlighted the importance of several areas, including the right MFG, right occipito-temporal gyri as well as bilateral posterior cerebellum, in facilitating typical sustained attention capacities in VPT adolescents, more particularly in a social context. Altogether, our findings indicate that despite preserved sustained attention competences at the behavioral level, VPT adolescents with increased socio-emotional weaknesses exhibit atypical recruitment of fronto-occipito-cerebellar regions compared to their FT counterparts, likely attributable to a more immature sustained attention cerebral network.

## Data Availability

All data produced in the present work are contained in the manuscript

## Acknowledgments

We thank and acknowledge all participating adolescents and their families who made this research possible. We also thank the Fondation Campus Biotech Geneva (FCBG), a foundation of the Ecole polytechnique fédérale de Lausanne (EPFL), the University of Geneva, and the University Hospitals of Geneva. Finally, we extend our gratitude to the paediatric clinical research Platform of the Geneva University Hospital for their help in data management.

## Funding

This work was supported by the Swiss National Science Foundation, No. 324730_163084 and the Von Meissner foundation.

## Notes

### Competing Interest Statement

The authors have declared no competing interest.

### Clinical Trial

ID: 215-00175

### Author Declarations

The Swiss Ethics Committees on research involving humans approved the all experimental protocols (ID: 215-00175). Written informed consent in accordance with regulations of the ethic committee at the University Hospital of Geneva was obtained from the principal caregiver and from the participant.

